# Small intestinal microbiota of undernourished women of reproductive age and microbiota-directed balanced energy protein (MD-BEP) supplementation in maternal environmental enteric dysfunction (EED): protocol for a community-based intervention study

**DOI:** 10.1101/2025.05.14.25327641

**Authors:** Md. Shabab Hossain, Mustafa Mahfuz, M Masudur Rahman, S.M. Khodeza Nahar Begum, Shafiqul Alam Sarker, Tahmeed Ahmed

**Affiliations:** Nutrition Research Division (NRD), International Centre for Diarrhoeal Disease Research, Bangladesh (icddr,b), Dhaka, Bangladesh; Department of Gastroenterology, Sheikh Russel National Gastro liver Institute & Hospital, Dhaka, Bangladesh; Department of Gastrointestinal, Hepatobiliary and Pancreatic Disorders, Bangladesh Specialized Hospital, Dhaka, Bangladesh; Department of Histopathology & Cytology, Bangladesh Specialized Hospital, Dhaka, Bangladesh; Department of Pathology, Dr Sirajul Islam Medical College, Dhaka, Bangladesh; Office of Executive Director (OED), International Centre for Diarrhoeal Disease Research, Bangladesh (icddr,b), Dhaka, Bangladesh; Department of Global Health, University of Washington, Seattle, Washington, United States of America

**Keywords:** Environmental enteric dysfunction, maternal undernutrition, microbiota directed balanced energy protein, gut microbiota, pregnancy

## Abstract

**Introduction:** Studies show, malnourished women of childbearing age with environmental enteric dysfunction (EED) exhibit small intestinal enteropathy resembling that in malnourished children residing in the same community. However, currently there are no universally accepted protocols for validation of these facts. Our current protocol is designed to better understand the mechanism of transmission of the microbiota of mothers with EED to their children perpetuates intergenerational undernutrition. We plan to compare the small intestinal (SI) and fecal microbiota along with plasma, duodenal, and fecal proteomes/ metabolomes and histopathologic evidence of EED in non-pregnant women with and without malnutrition. We also plan to see and compare the effect of microbiota-directed balanced energy protein (MD-BEP) supplementation on these biological parameters between malnourished non-pregnant and pregnant women.

**Methods and analysis:** This is a community-based intervention study where pregnant women and non-pregnant women of reproductive age (18-35 years) will be screened through household surveys from the Bauniabadh and adjacent slum area of Mirpur, Dhaka (pregnant and non-pregnant cohorts, n=90 each). Using Body Mass Index (BMI), both the groups will be categorized into undernourished (BMI <18.5kg/m^2^, n=60 each with pregnant and non-pregnant cohort) and well-nourished (BMI >20-24.9 kg/ m^2^, n=30, each with pregnant and non-pregnant cohort). Upper gastro-intestinal (UGI) endoscopy will be performed on non-pregnant women (both well-nourished and undernourished cohorts) and biopsy samples will be collected for diagnosis of environmental enteric dysfunction (EED) by histopathological scoring. We will compare the small intestinal (SI) and fecal microbiota and the plasma, duodenal, and fecal proteomes/ metabolomes of the undernourished non-pregnant cohort with histopathologic evidence of EED with the well-nourished non-pregnant cohort with no histopathologic evidence of EED (Aim IA). Furthermore, we will perform an intervention study (Aim IB). The undernourished non-pregnant cohort will be randomized into two groups (n=30/arm) and receive daily dietary supplementation with either shelf-stable microbiota-directed balanced energy protein (MD-BEP) or ready-to-use supplementary food balanced energy protein (RUSF-BEP) for 90 days. After cessation of the intervention they will be further followed up for another 270 days and biological samples will be collected at scheduled time points. The well-nourished non-pregnant cohort will not receive any nutritional intervention will serve as a reference control group. The undernourished pregnant cohort will also be randomly (n=30/arm) assigned to receive either MD-BEP or RUSF-BEP daily for 6 months until the child birth and thereafter for 3 months and followed up for another 9 months. During this period anthropometry will be measured and biological samples including fecal and plasma samples will be collected from the mothers and their infants in scheduled time points. Anthropometric, socio-demographic and laboratory assay data will be compared between the groups and candidate EED biomarkers will be correlated with nutritional status, histological analyses and score of EED, plus assessments of microbial community structure will be examined.

**Ethics and dissemination:** Ethics approval was obtained from the Ethical Review Committee of icddr,b (protocol no: PR-22117; Version 1.2; 29 November 2022). Results of this study will be submitted for publication in peer-reviewed journals.

**Trial registration number:** ClinicalTrials.gov ID: NCT05862363. Registered on 08 February 2023. https://clinicaltrials.gov/study/NCT05862363

## Introduction

Undernutrition among women of reproductive age is more common in South Asia than in any other region, with the prevalence of maternal undernutrition varying between 10% and 40%. Particularly in Bangladesh, the prevalence of undernutrition among women is much higher than in any other developing country, with more than 30% of women of reproductive age reported to be malnourished^1^. Maternal undernutrition has persistently been described to be a major contributor to child morbidity, mortality, and poor birth outcomes, including low birth weight (LBW), neonatal mortality, and subsequent childhood undernutrition. Maternal undernutrition alone accounts for about 25–50% of intrauterine growth restriction. In such a way, undernutrition can be transferred from one generation to other^1^. Half of the under-five children in the slums of Bangladesh are stunted with retardation of linear growth compared to one-third in non-slum areas^2^.

The prevention or treatment of intergenerational malnutrition represents a critical medical need yet to be addressed and remains a pressing global health challenge. Data are scarce on the contribution of small intestinal (SI) microbiota to pathogenesis, as it is difficult to obtain gut biopsy specimens from malnourished individuals. The Bangladesh Environmental Enteric Dysfunction (BEED) study, involving participants who live in an urban slum (Mirpur) in Dhaka, provided an opportunity to examine the role of the duodenal microbiota in the pathogenesis of environmental enteric dysfunction (EED) in children and its relationship to stunting^3^. 110 children, aged 18±2 months, who met the criteria for stunting or being at risk for stunting, failed to respond to nutritional intervention, and for whom consent was obtained, underwent esophagogastroduodenoscopy (EGD). The BEED study also performed EGD on thirty-eight 18-45-year-old malnourished (BMI<18.5 kg/m^2^) women residing in the same resource-poor setting of Mirpur, who failed to respond to an egg/milk/micronutrient-based nutritional intervention comparable to that given to children^3^. An operational categorization of EED was developed for this study based on three histopathologic features evident on hematoxylin and eosin-stained tissue sections; infiltration of inflammatory cells in the lamina propria, blunting or atrophy of small intestinal villi, and hyperplasia or elongation of crypts. The results, however, showed no statistically significant correlation between the length-for-age (LAZ) scores of the children.

Interestingly it was observed that 90% of the women had histopathologic evidence of enteropathy. The histopathologic features of their duodenal biopsies mimicked what was documented in the children with EED, including diminution in the height and number of villi, disruption of the small-intestinal (SI) epithelial barrier, and a chronic inflammatory infiltrate in the underlying lamina propria^4, 5, 6^. The study done by *Hossain et al* revealed that malnourished adults had a significantly higher frequency of subtotal villous atrophy, crypt hyperplasia, and marked cellular infiltration than the healthy controls^5^. Of the 38 women who underwent endoscopy, matched duodenal aspirates, fecal samples, and plasma samples from 22 [BMI 17.5±0.9 kg/m^2^; 26.6±8.2 years old] women were obtained. Matched sets of plasma samples and duodenal biopsies were collected from 84 of the 110 children. A ‘core group’ of 14 bacterial taxa were identified to be present in ≥80% of the EED-associated aspirates^4^. The absolute abundance of this shared group of 14 bacterial strains, recovered from the duodenums of children with histopathologic evidence of EED and not typically classified as enteropathogens, were found to be negatively correlated with linear growth, and positively correlated with components of the expressed duodenal proteome involved in immuno-inflammatory responses.

Based on these findings, in this proposal, we will test the hypothesis that small intestinal microbiota contributes to small intestinal enteropathy and malnutrition in young Bangladeshi women of childbearing age. Based on the hypothesis that transmission of microbiota of mothers with EED to their children perpetuates intergenerational undernutrition, an interventional component will be included involving the administration of a specially designed nutritional supplement aimed at improving gut microbiota in pregnant and non-pregnant low BMI women, termed as the microbiota directed balanced energy protein (MD-BEP).

### Nutritional intervention

Conventional nutritional interventions or low-cost water, sanitation and hygiene (WASH) interventions were found ineffective in reversing EED-related malnutrition^7–9^, warranting microbiome-targeted food and other interventions. Prototypes for nutritional interventions composed of locally available, affordable, culturally acceptable complementary foods commonly consumed in Bangladesh have recently been developed. Preclinical studies using gnotobiotic mice and piglets colonized with members of the gut microbiota from Bangladeshi children with acute malnutrition revealed that these formulations contain nutrients that increase the representation and expressed beneficial functions of growth-promoting gut bacterial strains that are underrepresented in the microbiota of affected children. Several of these microbiota-directed complementary food (MDCF) formulations were subsequently tested in a pre-POC study involving 12–18-month-old Bangladeshi children with moderate acute malnutrition (MAM) living in the same slum (Mirpur). This 1-month long, four-arm, controlled feeding study tested three MDCFs and a commonly used, rice-lentil-based, ready-to-use supplementary food (RUSF) that was not designed to change gut microbial community structure or function. One of the MDCFs, MDCF-2, was distinguished from the other formulations based on its superior ability to (i) repair the microbiota of children with MAM to a configuration that resembled that of healthy individuals living in the same community, and (ii) change the levels of multiple plasma proteins involved in mediating various aspects of metabolism, bone growth, immune function and neurodevelopment towards a healthy state^10, 11^. These results support the notion that repair of impaired gut microbial community development could represent a new therapeutic concept for restoring healthy growth^10^. MDCF-2 is composed of chickpea flour, peanut flour, soy flour, green banana, sugar, soybean oil, and vitamin-mineral premix. Based on the findings, working with a large food manufacturer of Bangladesh, a shelf-stable, packaged formulation of MDCF was developed, which was named MDCF shelf-stable foil pouch formulation with green banana powder.

The new WHO antenatal care guideline has made a recommendation of balanced energy protein (BEP) supplementation for pregnant women in undernourished populations to reduce the risk of stillbirths and small-for-gestational-age neonates. Several recent meta-analyses also showed that BEP supplementation during pregnancy had a pronounced positive effect on infant birth weight among women in undernourished settings^12–14^. The Bill and Melinda Gates Foundation organized an expert consultation in September 2016 for the purpose of developing nutrient content targets for affordable, nutritional supplements for use by pregnant and lactating women in undernourished settings. The expert consultation examined the different types of formulations that were used in previous BEP trials and considered the Dietary reference intakes (DRIs) of macro and micronutrients for pregnant and lactating women proposed by the U.S. Institute of Medicine (IOM) and FAO/WHO. The expert consultation recommended that women in undernourished settings should receive a daily serving of BEP supplement containing 250-500 kcal of energy and 14-18 grams of protein. In a high-risk population, when large protein and energy gaps exist, the portion size could be doubled. Alternately, in a low-to-moderate risk context, the supplement could provide the lower daily energy value^15^. Based on the recommendations, in this study, we developed a modified BEP formulation with the ingredients used in MDCF-2, which is aimed to improve the gut microbiota and are calling it microbiota directed balanced energy protein (MD-BEP). The other arm will receive another modified BEP developed with the ingredients used in RUSF and are calling it ready-to-use supplementary food-balanced energy protein (RUSF-BEP).

### Hypothesis and Objectives

Our previous studies suggest that malnourished women of childbearing age living in Mirpur exhibit small intestinal enteropathy resembling that found in Mirpur children with EED^4,5,6,18^. In our current study, we will test the hypothesis that SI microbiota contributes to small intestinal enteropathy and malnutrition in young Bangladeshi women of childbearing age. An interventional component will be included involving the administration of MD-BEP or RUSF-BEP in pregnant and non-pregnant undernourished women. This will be based on the hypothesis that transmission of the microbiota of mothers with EED to their children perpetuates intergenerational undernutrition. The overarching goals for the current study will be to (i) delineate mechanisms by which the SI microbial community obtained from undernourished Mirpur women contributes to maternal malnutrition and identify surrogate biomarkers that can be applied to malnourished pregnant women, and (ii) test whether MD-BEP can ameliorate EED as judged by these surrogate endpoints in undernourished women (who are either pregnant or non-pregnant).

## Methods and analysis

This is a community-based intervention study where participants will be enrolled in two cohorts: undernourished pregnant (BMI <18.5kg/m^2^; 18-35 years, n=60) and undernourished non-pregnant ((BMI <18.5kg/m^2^; 18-35 years, n=60) women from Bauniabadh and adjacent slum areas of Mirpur, Dhaka. For comparison, well-nourished pregnant women (BMI 20-24.9kg/m^2^; 18-35 years, n=30), who will be screened through household surveys from the Bauniabadh and adjacent slum area of Mirpur, Dhaka, and well-nourished non-pregnant (BMI 20-24.9kg/m^2^; 18-35 years, n=30) women of reproductive age will be recruited. We will use two specific aims, AIM 1A and AIM 1B. In AIM 1A, we will compare the small intestinal (SI) and fecal microbiota and the plasma, duodenal, and fecal proteomes/ metabolomes of the undernourished non-pregnant cohort with histopathologic evidence of EED with the well-nourished non-pregnant cohort with no histopathologic evidence of EED undergoing routine endoscopic evaluation for dyspepsia. Upper gastro-intestinal (UGI) endoscopy will be performed on non-pregnant women and biopsy samples will be collected for diagnosis of EED and histopathological scoring. In AIM 1B, we will perform an intervention study. After UGI endoscopy, undernourished non-pregnant cohort will be randomized into two groups (n=30/arm) and receive daily dietary supplementation with either shelf-stable microbiota-directed balanced energy protein (MD-BEP) or ready-to-use supplementary food balanced energy protein (RUSF-BEP) for 90 days. After cessation of the intervention they will be further followed up for another 270 days and biological samples will be collected at scheduled time points. The well-nourished non-pregnant cohort will not receive any nutritional intervention will serve as a reference control group. The undernourished pregnant cohort will also be randomly (n=30/arm) assigned to receive either MD-BEP or RUSF-BEP daily for 6 months until the child birth and thereafter for 3 months and followed up for another 9 months. During this period anthropometry will be measured and biological samples including fecal and plasma samples will be collected from the mothers and their infants in scheduled time points. The well-nourished pregnant cohort will not be supplemented with MD-BEF or RUSF-BEP but will be followed-up with anthropometric measurement and sample collection like undernourished pregnant cohort. Due to ethical concerns related to performing endoscopy in pregnant women, UGI endoscopy in pregnant women will not be performed. Anthropometric, socio-demographic and laboratory assay data will be compared between the groups and candidate EED biomarkers will be correlated with nutritional status, the results of histological analyses, plus assessments of microbial community structure will be examined. After complete disclosure, a signed written informed consent statement is obtained from each subject. If the subject agrees to participate in the study, they sign the consent form or if unable to sign, provide an impression of their thumb. For endoscopy and supplement intervention, a separate consent form is used and the aforementioned procedure is followed. The consent form for endoscopy clearly and fully describes, and demystifies, all aspects of the process, including the risks related with the procedure. No information is remained withheld from the participant. While enrolling the neonates, a signed written informed consent from the parents or guardians is obtained. All consents are obtained and documented in presence of the study physician at the study field office in front of a witness, who also sign the document as the witness. The first participant from the non-intervention arm was enrolled on 28 April, 2023, while the first participant from the intervention arm was enrolled on 09 May, 2024. Recruitment is currently ongoing and the tentative end date for completing the recruitment is 31 October, 2025. The complete data collection (including one-year follow-up data) is estimated to be completed by 31 October, 2026. Results are expected by 31 December, 2026. The schedule of study events is presented in Fig. 1.

**Fig. 1.**
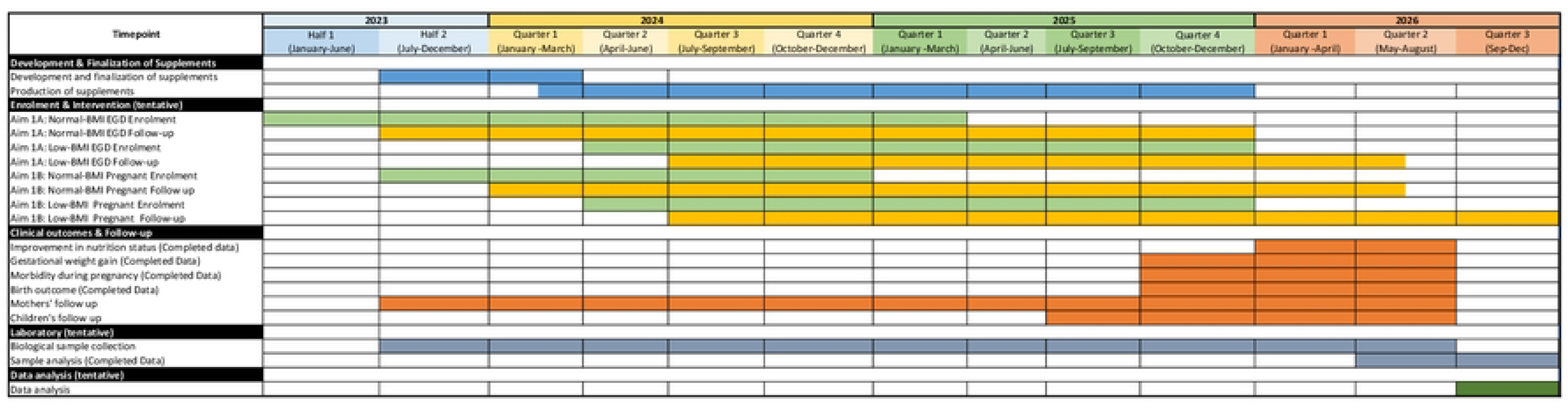

### AIM 1A: Non-pregnant cohort

In AIM 1A, for the healthy group, we plan to recruit a cohort of well-nourished Bangladeshi women who will undergo esophagogastroduodenoscopy (EGD) for evaluation of functional dyspepsia and identify 30 well-nourished participants who have normal duodenal mucosal histology. Healthy women (BMI 20-24.9 kg/m^2^) of childbearing age who have been referred for evaluation of functional dyspepsia as defined by Rome IV criteria, will be enrolled from women attending the Gastroenterology OPD of Bangladesh Specialized Hospital (BSH), Dhaka, Bangladesh, and also from the community who meet the inclusion criteria. Duodenal biopsies, duodenal aspirates, plasma and fecal specimens from these participants at the time of endoscopy and serial plasma and fecal samples will be collected according to the follow-up schedule (Fig.1). These women will be considered as the control group for the non-pregnant cohort and will be followed for 360 days but will not receive any nutritional intervention. Undernourished (<18.5kg/m^2^; 18-35 years) women of childbearing age will be enrolled from Bauniabadh and adjacent slum areas of Mirpur, Dhaka, and EGD will be performed among 60 women at icddr,b Dhaka Hospital, or BSH, Dhaka. Participants will be screened through household surveys from the Bauniabadh and adjacent slum area of Mirpur, Dhaka. After EGD, the undernourished women will be randomized into two groups (n=30/arm) and receive daily dietary supplementation with either MD-BEP or RUSF-BEP for a period of 90 days, with a further 270 days of follow-up after cessation of the intervention, and biological samples will be collected from the participants according to the schedule. The EGD scheme is presented in Fig. 2.

**Fig. 2.**
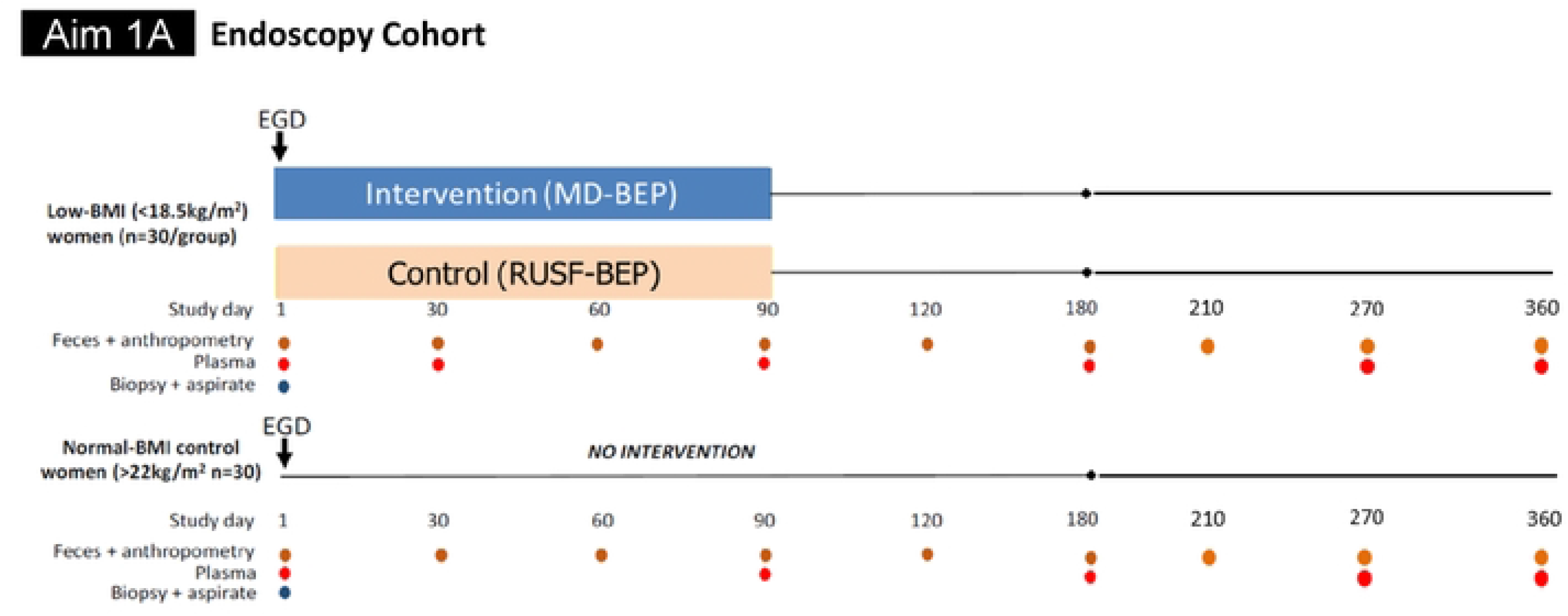

### AIM 1B: Pregnant cohort

For AIM 1B, beginning at the end of the first trimester, undernourished (<18.5 kg/m^2^) pregnant women (aged 18-35 years) will be randomly assigned to receive either MD-BEP or RUSF-BEP for the duration of their pregnancy and during the first three postnatal months, in addition to standard antenatal care (n=30/arm). A parallel cohort of age-matched well-nourished pregnant women (n=30) who will not receive any nutritional intervention will serve as a reference control group. Due to safety, sensitivity, and ethical concerns related to performing endoscopy in pregnant women, EGD will not be performed on these women. The women will receive standard antenatal care, will be followed up and serial plasma and fecal samples will be collected according to Fig. 3. These women will be considered as the control group for the pregnant cohort and will not receive any nutritional intervention.

**Fig. 3.**
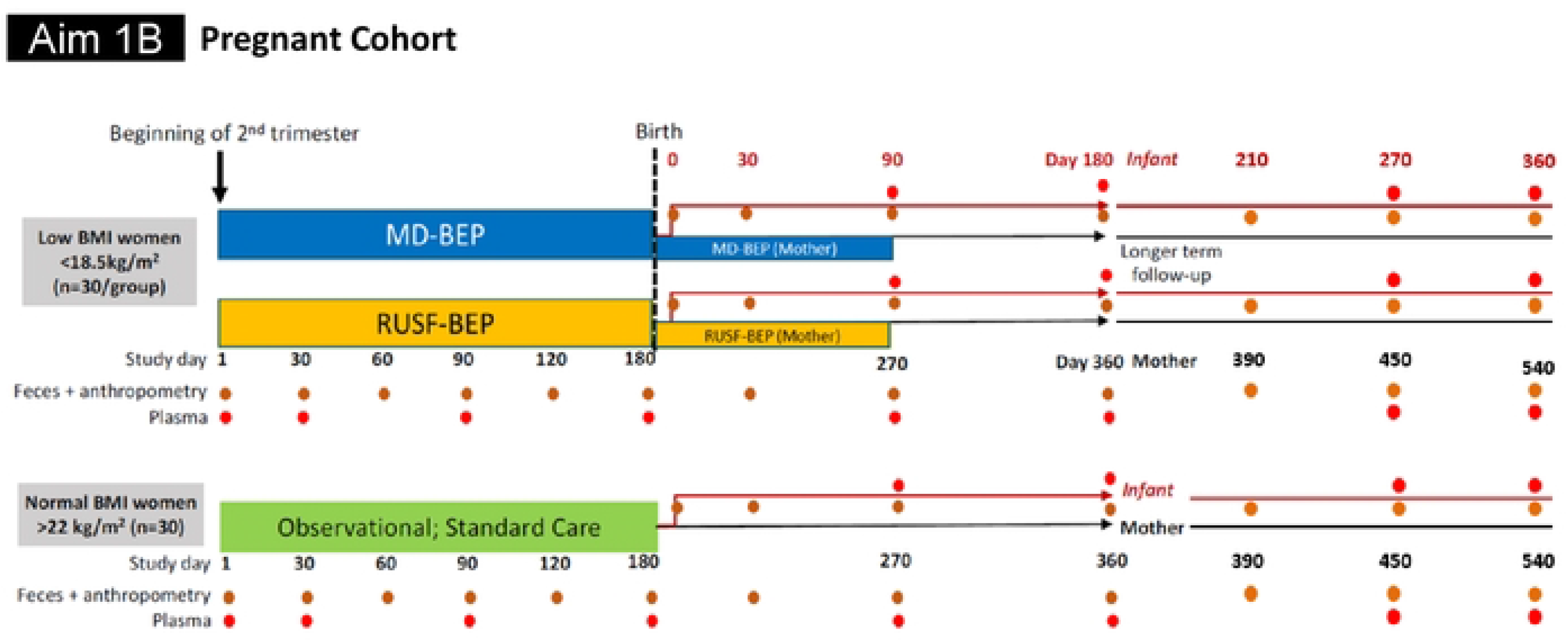

### EGD and biopsy sample collection

Endoscopy will be performed at icddr,b Dhaka Hospital and BSH. Clinical metadata will be collected, including socio-demographic data, dietary history, and use of antibiotics and proton pump inhibitors (PPI) during 12 months prior to EGD. EGD will be performed according to standards recommended by the American College of Gastroenterology (ACG) and the American Society for Gastrointestinal Endoscopy (ASGE)^16^. The endoscopist will use sterile ERCP catheters to collect secretions from the second portion of the duodenum (prior to biopsy). Aspirates (~1-2 mL/subject) will be divided into aliquots and stored in a manner that preserves bacterial viability using the protocol we employed in the BEED study for children with EED^4^.

### Histopathology definition of EED

After EGD, histopathology will be performed to identify women who have EED. An operational categorization of EED used previously based on three histopathologic features (infiltration of inflammatory cells in the lamina propria, blunting or atrophy of small intestinal villi, and hyperplasia or elongation of crypts of Lieberkuhn) evident in hematoxylin- and eosin-stained tissue sections will be employed^17^. Histopathologic and immunocytochemical analyses will be conducted at BSH. The biopsies will also be processed at BSH. The standard operating procedure used at Bangladesh Specialized Hospital (BSH) will be followed. The scoring systems developed by *Liu et al* and the categorization of intestinal histomorphology by *Hossain et al* will be used for the H&E sections ^5,6,17^. Because inflammatory infiltration takes place before structural changes of either villus atrophy or crypt hyperplasia-

▪ Mild EED (Grade 1) was defined as the sole presence of inflammatory infiltrates in the lamina propria.
▪ Moderate EED (Grade 2) indicated the presence of one of two structural changes (either villi or crypts) in addition to inflammatory infiltration in the same biopsy sample.
▪ Severe EED (Grade 3) was defined based on the presence of structural changes involving both villi and crypts in addition to inflammatory infiltration in the same biopsy sample.
▪ Biopsies without these three features were considered as having no evidence of EED (Grade 0).

### Processing of collected biological samples

Schedule for biological sample collection is indicated in Figs 1 and 2. All biological samples (blood and stool) will be collected as per standard operative procedures (SOPs) prepared for this protocol. A total amount of 5 ml blood will be collected from each participant. Collected fecal samples will be cryopreserved within 20 minutes of defecation. At the site of collection, samples will be aliquoted into sterile, pre-labelled 2 mL cryovials and immediately placed into pre-charged liquid nitrogen dry cryo-shippers for transport back to the laboratory. At the laboratory, vials will be transferred from the dry shipper to a bucket of dry ice while they are organized and transferred to 9×9 freezer boxes and immediately placed into a −80°C freezer. No additives, preservatives or media will be added to the fecal samples. The preparation and transport of all biological samples will strictly adhere to the biosafety laws and procedures of both institution’s and international standards. An aliquot of each fecal sample will be used at icddr,b for (i) Taqman Array Card (TAC)-based assays of 34 enteropathogens and (ii) commonly used clinical markers of intestinal inflammation and barrier disruption [ELISA of several fecal (and plasma) biomarkers including CRP, AGP, myeloperoxidase, neopterin, alpha-1 anti-trypsin, calprotectin, Reg1B, lipocalin-2 (LCN2), and Duox-2].

### Details of intervention products

#### Microbiota directed-balanced energy protein (MD-BEP)

Every woman in the MD-BEP arm will be offered a single serving of MD-BEP daily, aimed as a supplement to the usual diet received by these women. The feeding will initially take place in the field office under direct supervision for at least one week, and continued at home afterwards. The feeding will be supervised by the respective field research assistants (FRAs) in regular intervals at home. MD-BEP will provide approximately 500 Kcal, 14 g of protein and the necessary micronutrients, and the substrate for the beneficial microbiota proliferation.

#### Ready-to-use supplementary food-balanced energy protein (RUSF-BEP)

Women in the RUSF-BEP group will receive a single serving of RUSF-BEP daily, similar to the MD-BEP arm. The feeding will initially in the field office for at least one week, and continued at home afterwards. The feeding will be supervised by the respective FRAs. Each serving of RUSF-BEP will also provide approximately 500 Kcal, 14 g of protein, and necessary micronutrients following the recommendations from the expert panel convened by the BMGF on nutritional supplementation in pregnancy^15^.

The amount in each serving of the supplements and the compositions are shown in tables 1 and 2, respectively, which are in conformity with the recommendations from the expert panel convened by the BMGF on nutritional supplementation in pregnancy^15^.

**Table 1.**
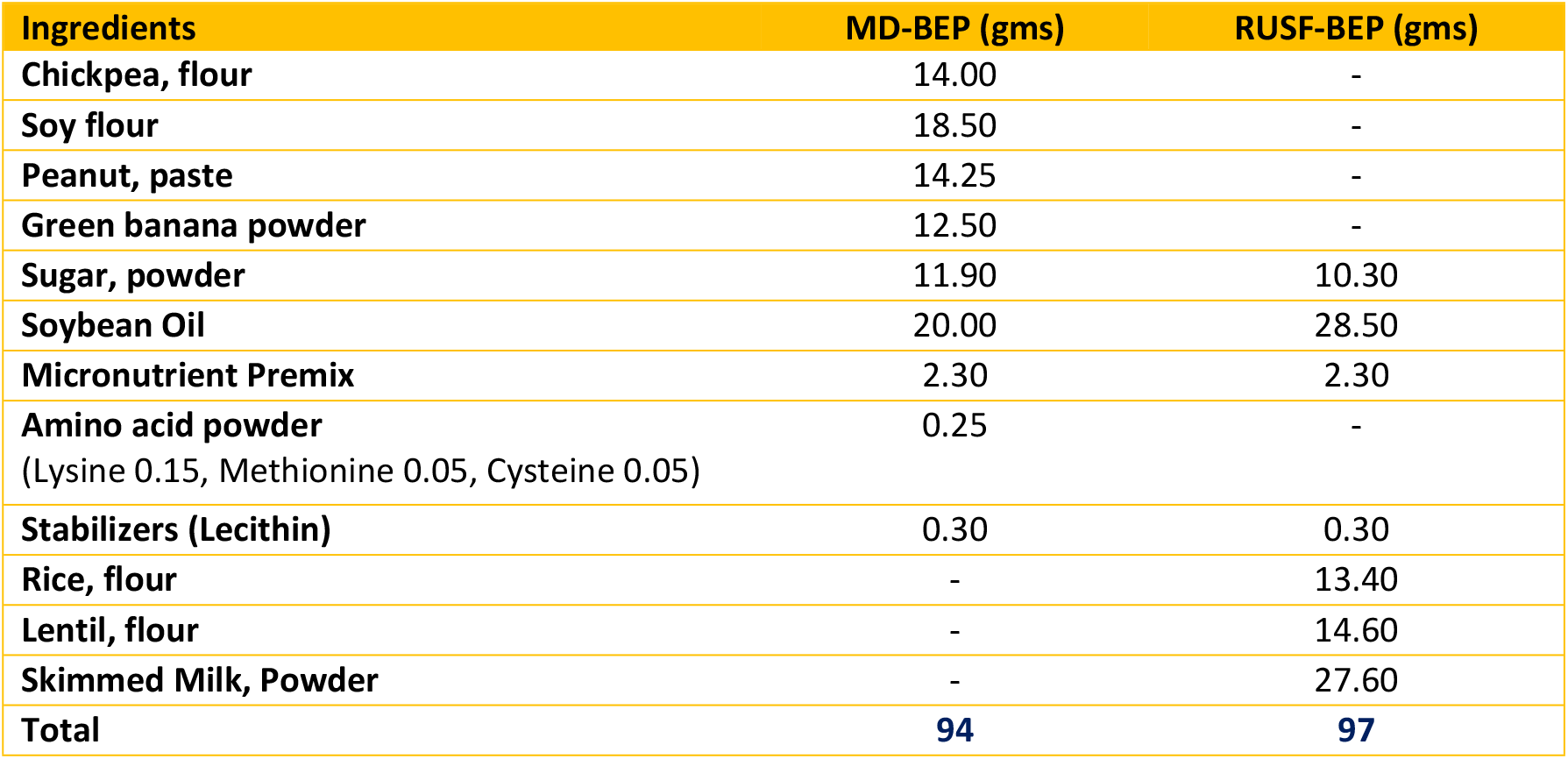
Ingredients used in MD-BEP and RUSF-BEP formulations with amount per serving.

| Ingredients | MD-BEP (gms) | RUSF-BEP (gms) |
| --- | --- | --- |
| Chickpea, flour | 14.00 | - |
| Soy flour | 18.50 | - |
| Peanut, paste | 14.25 | - |
| Green banana powder | 12.50 | - |
| Sugar, powder | 11.90 | 10.30 |
| Soybean Oil | 20.00 | 28.50 |
| Micronutrient Premix | 2.30 | 2.30 |
| Amino acid powder<br>(Lysine 0.15, Methionine 0.05, Cysteine 0.05) | 0.25 | - |
| Stabilizers (Lecithin) | 0.30 | 0.30 |
| Rice, flour | - | 13.40 |
| Lentil, flour | - | 14.60 |
| Skimmed Milk, Powder | - | 27.60 |
| **Total** | **94** | **97** |

**Table 2.** Nutritional composition of MD-BEP and RUSF-BEP.

| Nutritional composition | BEP recommendation | MD-BEP<br>(per 94g) | RUSF-BEP<br>(per 97g) |
| --- | --- | --- | --- |
| Total Energy(kcal) | 250-500 | 494.62 | 497.54 |
| Protein (g) | 14-18 | 14.18 | 14.33 |
| Energy from Added Sugar,<br>(% of total energy) | <10% | 9.36% | 8.05% |
| Energy from Fat (FER),<br>(% of total energy) | 10-60% | 58.48% | 53.08% |
| Total ω-3 (g) | $\geq 1.30$ | 1.70 | 2.01 |
| (ω-6/ω-3) ratio | 5-10:1 | 8.29:1 | 7.34:1 |
| Total Trans-fatty acid (g) | <1.00 | 0.16 | 0.19 |
| DIAAS | $\geq 90\%$ | 91.67% | 123.64% |

The enrolled women will be monitored regularly by Field Research Assistants for any possible side effects/adverse events (e.g. rash, urticaria due to food allergy, or any significant changes in clinical status) for a week. If any side effects/adverse events are observed, they will be treated according to the standard of care. Study participants will be given dietary counselling with particular emphasis on ensuring dietary diversity, adding vegetable oil to the cooked diet as a source of energy, as well as sources of animal-based products such as small fish or chicken meat etc. along with regular intake of vegetables. The study participants will also receive support for any intercurrent illnesses detected by the study team as well as proper referral will be provided.

#### Manufacturing of intervention products

The servings will be provided in sterilized formulations packaged in trilaminar foil pouches developed in Natore Agro Limited (NAL) factory, a sister concern of a renowned food manufacturing company of Bangladesh, PRAN. A standardized production process will be followed to maintain the quality of the formulations throughout the project period. Raw materials will be at first passed through a vigorous quality-checking process. The materials will then be roasted in a temperature-controlled hot air oven, followed by a grinding process using a heavy-duty electric grinder to make fine powder/paste as appropriate. After mixing, the mixture will be measured into the trilaminar foil pouches. Each step of the food processing will be closely monitored. An Impulse Sealer Machine is used to achieve airtight sealing of the pouches. Sterilization is performed in a retort machine at 105°C and 0.123 MPa pressure for 10 minutes. This process will ensure a longer contamination-free shelf life while preserving the organoleptic properties of the food. Before finalization, the supplement prototypes will undergo microbiological safety and organoleptic test for acceptable results. During manufacturing and distribution to the study site, random samples from each batch will be tested for organoleptic properties and microbial contamination.

#### Data collection

At the beginning of the study, information will be sought on the demographic characteristics (wealth, husband’s occupation and income, standard of housing, family structure, and home environment etc.). Morbidity data will be collected weekly during the intervention period.

#### Anthropometry and body composition

Nutritional status will be assessed through anthropometry. Body weight and BMI will be determined at the anthropometry timepoints mentioned in Figs 1 and 2. Bioelectric Impedance Analysis (BIA) will be used to measure total fat and fat-free mass before, during and after the intervention.

#### Follow-up of pregnant women

Pregnant women will be enrolled at the beginning of second trimester and followed up till one year after childbirth. For the undernourished pregnant women, antenatal care (ANC) services from nearby healthcare facilities will be ensured by study staff. Trained Health Workers (HWs) will visit the homes of all enrolled pregnant women once a month and will involve the family members, including mothers, mothers-in-law, and husbands, along with the pregnant women in the sessions according to the convenience and availability of the participant and her family members. The woman and her family will receive advice on the importance of adequate weight gain during pregnancy, additional energy requirements, and improving dietary diversity. The pregnant women will be advised to seek routine ANC according to the schedule and to regularly take micronutrients (iron, folic acid, and calcium) provided through routine ANC. On each visit, the participants’ dietary diversity and rate of weight gain will be evaluated, and they will be given instant feedback. The participants will have access to field offices where they shall receive a thorough physical check-up by a registered physician. If any unwanted event occurs during the follow-up period, the participants will be referred to the nearest maternal clinic or appropriate healthcare facility.

#### Available health care services for low-BMI pregnant women in the study site

The Bauniabadh area of Dhaka city is located in ward-5 of Dhaka North City Corporation^3,18,19^. This urban area has availability of different facilities and services for secondary and tertiary-level health care. Medical clinics and diagnostic centers have become numerous all over the area outside slums. In this area, health infrastructure facilities owned and operated by different entities can be categorized into (i) public, (ii) private, and (iii) NGOs. Currently, in this particular area, primary health care targeting maternal and child nutrition is provided by the ADB-supported Urban Primary Health Care Project (UPHCP II), the USAID-supported NGO Services Delivery Program, which preceded the ongoing Smiling Sun Franchise Program (SSFP), Urban Health Center by BRAC, and Radda MCH-FP center. The services provided by these facilities include basic medical consultation, diagnostics, EPI, pregnancy and delivery services, including ANC, Infant and Young Child Feeding (IYCF) services, and nutrition counseling.

#### Sample size calculation and outcome

For normal-BMI (BMI 20-24.9 kg/m^2^) women of childbearing age or healthy volunteers undergoing EGD, the goal is to identify 30 normal-BMI participants who will undergo EGD for evaluation of functional dyspepsia and will have normal duodenal mucosal histology. We plan to recruit 100 individuals for EGD, and anticipate that 60 will have duodenal mucosa without macroscopic evidence of active acid-peptic disease, of whom 30 will have normal duodenal mucosal histology. These calculations are based on the team’s gastroenterologist who has 15-year experience with >30,000 subjects from this socioeconomic group and geographic locale. These specimens from normal BMI participants without enteropathy will be compared with those of 60 undernourished low-BMI (<18.5kg/m2) women of childbearing age. As this is an experimental study, no formal power calculation has been performed for this exploratory research study. The same applies for the pregnant cohort.

### Data analysis

The cohort-specific characteristics of the recruited participants will be described by exploratory data analysis. Normally distributed variables will be summarized as mean and standard deviation, whereas, median and inter-quartile range will be used for the variables following skewed distributions. Binary and categorical variables will be presented as counts and percentages. Appropriate statistical tests (Student’s t-tests, Pearson chi-square tests, and Mann-Whitney test) will be used to detect the differences in baseline characteristics of the two intervention groups. A probability of <0.05 will be considered statistically significant. The correlation between the confounders and outcome will be evaluated using Pearson or Spearman correlation, whichever is appropriate. Multi-collinearity between the predictor variables will be assessed using variance inflation factor (VIF), and a predefined cut-off of 2 will indicate the absence of multi-collinearity.

#### Modeling the clinical effectiveness of the intervention

Linear mixed effect model - a longitudinal data analysis technique will be used to measure the role of intervention on the responses listed above. The mixed effects model for continuous outcome data will be presented as Yi = Xi β + Zibi + εi, where Yi is the vector of responses for the subject over the follow-up time points, Xi is the matrix for the fixed effects (treatment, baseline covariates, average time trends, etc.) with corresponding regression coefficients β, Zi is the matrix for the random effects of individual subject trends (morbidity, adherence to the intervention, etc.) with corresponding coefficients b, and εi is the vector of residuals. Both intention-to-treat and per-protocol analyses will be performed.

#### Modeling the changes in centiles of anthropometry curves

Percent changes in weight and BMI will be calculated by deducting the values from the baseline, dividing the outcome by the baseline values, and multiplying by 100. Generalized linear mixed-effects models (GLMMs) will be used to model the percent changes of indices. GLMMs will estimate the probability of improvement in the BMI and weight in the intervention group from baseline in comparison to the control group, after adjusting for all the other possible covariates.

### Data Safety Monitoring Plan (DSMP)

Data collection tools for this study will include case report forms, laboratory worksheets and source documentation. Complete source documentation (study visits, laboratory reports, etc.) will be kept for each participant in individual study charts. All biological samples will be collected following the Good Clinical Practice (GCP) guidelines. All laboratory specimens, reports, study data collection, and administrative forms will be identified by coded number to maintain participant confidentiality and to enable tracking throughout the study. All study related documents will be kept in locked cabinets in locked rooms with limited access. Information in the electronic database established at icddr,b will be password-protected and access will be available only to authorized research team members. Data entry and cleaning will be done at icddr,b.

### Adverse events

Expected AEs for this protocol are those related to the endoscopy/biopsy procedure, related to pregnancy and childbirth, and those associated with phlebotomy. Both serious and non-SAEs are assessed for their severity, their relationship to study participation and the actions taken and their outcomes. All SAEs are being reported to the ERC of icddr,b within 24 hours of the site’s awareness of the event. In the event that medical care is required outside of the protocol, all necessary and available treatments are provided or proper referrals are ensured.

## Data Availability

No datasets were generated or analysed during the current study. All relevant data from this study will be made available upon study completion.

## Ethics and dissemination

### Ethics approval and consent to participate

#### Ethical approvals

Ethical Review Committee of icddr,b (protocol no: PR-22117; Version 1.2; 29 November 2022)

#### Consent

Each participant enrolled in the study is treated according to what is morally right and proper. After complete disclosure, a signed written informed consent statement is obtained from each subject. If the subject agrees to participate in the study, they sign the consent form or provide an impression of their thumb. For endoscopy and supplement intervention, a separate consent form is used and the aforementioned procedure is followed. The consent form for endoscopy clearly and fully describes, and demystifies, all aspects of the process, including the risks related with the procedure. No information is remained withheld from the participant.

#### Dissemination and publication

The data, results and other findings resulting from this study will be published only after approval by a committee consisting of the investigators of the protocol. The International Committee of Medical Journal Editors guidelines will be used to establish authorship on papers.

## Acknowledgements

This work was supported, in whole, by the Gates Foundation (https://www.gatesfoundation.org/about/committed-grants/2022/04/inv045337). Project investment ID is INV-045337.The conclusions and opinions expressed in this work are those of the author(s) alone and shall not be attributed to the Foundation. Under the grant conditions of the Foundation, a Creative Commons Attribution 4.0 License has already been assigned to the Author Accepted Manuscript version that might arise from this submission. Please note works submitted as a preprint have not undergone a peer review process. icddr,b acknowledges with gratitude the commitment of Bangladesh Specialized Hospital, Dhaka to its research efforts. icddr,b also gratefully acknowledges the following donors who provide unrestricted support: Government of the People’s Republic of Bangladesh and Global Affairs Canada (GAC).

## Author contributions

TA originated the idea for the study and led the protocol design. TA, MM and MSH participated in the design of the study. TA, MM, MSH, MMR, SAS, and SMKNB were involved in the development of the study protocol. MSH, MM, and TA were involved in the implementation. MM, MSH and TA were involved in the data analysis plan. All authors read and approved the final manuscript.

## Project status

Participant enrolment is ongoing. Running protocol version v1.3, dated 07 May 2024. Participant recruitment commenced on 15 April, 2023 and estimated end date is 30 September 2026. As of May 2025, participant enrolment is ongoing.

